# Emotional responses and coping strategies of nurses and nursing college students during COVID-19 outbreak

**DOI:** 10.1101/2020.03.05.20031898

**Authors:** Long Huang, Fuming Xu, Hairong Liu

## Abstract

**Background:** Affected by a Corona Virus Disease 2019 (COVID-19) outbreak, Since December 2019, there have been more than 76,000 cases of COVID-19 in China, causing more than 3,000 medical staff infections. Due to COVID-19 spreads quickly, is highly contagious, and can be fatal in severe cases, and there are no specific medicines, it poses a huge threat to the life and health of nurses and has a large impact on their emotional responses and coping strategies.

**Methods:** This study conducted an online questionnaire survey from February 1 to 9, 2020 to investigate the current state of emotional responses and coping strategies of nurses and college nursing students in Anhui Province. This study used a modified Brief COPE (Carver, 1997) and a emotional responses scale.

**Results:** The results found that women showed more severe anxiety and fear than men. Participants from cities showed more anxiety and fear than participants from rural, but rural participants showed more sadness than urban participants. The closer COVID-19 is to the participants, the stronger the anxiety and anger. Compared with Nursing college students, nurses have stronger emotional responses and are more willing to use Problem-focused coping. People may have a cycle of “the more fear, the more problem-focused coping”. And people may “The more angry, the more emotion-focused coping”, “the more problem-focused coping, the more anxious, the more angry, the more sadness”.

**Conclusion:** COVID-19 is a pressure source with great influence, both for individuals and for the social public groups. Different individuals and groups may experience different levels of psychological crisis, and those nurses at the core of the incident are affected. Hospitals should focus on providing psychological support to nurses and providing timely psychological assistance and training in coping strategies.Improving nurses’ ability to regulate emotions and effective coping strategies, providing a strong guarantee for resolutely winning the battle against epidemic prevention and control.

## Introduction

On December, 2019, Multiple unexplained cases of pneumonia were reported in Wuhan, Hubei Province, China. Epidemiological findings revealed severe human-to-human transmission, which was later confirmed to be caused by a novel corona virus (2019-nCoV) infection. The World Health Organization (WHO) named it Corona Virus Disease 2019 (COVID-19) [1]. Although 13 prefecture-level cities in Hubei, including Wuhan, and one autonomous prefecture have announced closures from January 23, 2020. The Chinese government also announced the extension of the Spring Festival holiday (from 7 days to 14 days), and implemented epidemic prevention and control measures such as suspension of school, shutdown, and closure of business to reduce the concentration of personnel and block the spread of the epidemic. However, due to the large population migration before the Chinese Lunar New Year, more than 5 million residents have reportedly left Wuhan for various cities in China and around the world. Compared with viruses such as SARS and Ebola, COVID-19 is highly infectious during the incubation period, and asymptomatic infection exists. It can be transmitted through respiratory droplets, contact and aerosols [2]. As a result, large-scale infection of COVID-19 has been caused worldwide. As of February 22, 2020, China has reported 51,699 confirmed cases (including Hong Kong, Macao, and Taiwan, including 10,968 severe cases), a total of 22,888 discharged patients, 2442 deaths, and 76,936 confirmed cases. There were 4148 suspected cases. A total of 628,517 close contacts were tracked, and 106,089 close contacts were still in medical observation. A total of 1,719 confirmed cases and 17 deaths have occurred in other countries around the world.

Due to the rapid spread of COVID-19, strong contagion, lethality in severe cases, and no specific medicine, it poses a huge threat to human life and health, and also has a huge impact on the mental health of the general public, causing people to differ degree of emotional problom [3]. 17 years ago, the emergence of SARS in China also caused widespread serious concerns, fears and heightened emotions [4,5]. So we can foresee that the outbreak of COVID-19 will course the public psychological reactions such as tension, anxiety, and fear which will lead to psychological disorders such as acute stress disorder post-traumatic stress disorder, depression and suicide. Academician Zhong Nanshan, the leader of the high-level expert group of the Chinese Health Commission, pointed out that psychological fear is more fearful than the disease itself [6]. Although infectious diseases elicits a wide range of emotional responses, not everyone experiences the same degree of emotional impact [7]. Hospital medical staff is always at the forefront of any particular epidemic and they risk their lives to perform their duties. Because they are more likely to be in close contact with COVID-19 patients, they are particularly vulnerable to infection and spread the virus among colleagues and family members. To date, more than 3,000 medical staff have been infected with COVID-19, and six of them have died. The number of infected medical personnel is unique in modern history. In addition to physical stress, medical staff also face huge mental burdens. This is particularly evident in the SARS and Ebola virus outbreaks [8,9]. Previou studies have found that SARS causes great pain to medical staff, and it causes much more pain to nurses than doctors [10]. This is due to the nature of the nurse’s job, which makes it easier to stay with patients for a long time. In addition, many hospital temporary workers resigned during the outbreak, and a large part of their work could only be performed by nurses. Therefore, the emotional problems of nurses during the COVID-19 epidemic deserve more attention.

In addition to the impact of the COVID-19 on people’s emotions, people’s coping strategies will also change as a result. Coping is the thoughts and actions that individuals use to deal with stressful events [11]. People have identified two general coping strategies: one is a problem-focused coping, the purpose is to solve the problem or take action to change the status quo; the other is an emotion-focused coping, which aims to reduce the emotional distress associated with stressful situations [12]. Studies have found that emotions lead to specific coping strategies [13], and vice versa. Emotions are thought to have motivational properties that motivate certain behaviors or behaviors [13,14]. For example, fear is related to the desire to evade and protect themselves from incidents, anger leads to desire to attack, disgust leads to desire to expel, and happiness leads to desire to entertain [13,15]. Moreover, emotions have been linked to the use of specific coping strategies [16]. In particular, adults who report more anger and fear prefer to use active-oriented coping strategies such as asking questions, while those who are sad are more likely to use non-active coping strategies such as avoiding or accepting problems [16]. In turn, the successful use of coping strategies will help individuals manage stressful events [11] and reduce negative emotions [12]. However, the direction of the relationship between emotion responses and coping strategies is not clear, and the relationship is not always constant. Some studies have found that the relationship between them is age-specific during SARS [17]. So, the relationship between nurses’ coping strategies and emotional response during a major infectious disease such as COVID-19 needs further research to clarify. To our knowledge, there has been no systematic assessment of the effects of COVID-19 on nurses’ emotional responses and coping strategies. Based on this, the purpose of this study was to explore the current status and relationship of emotional responses and coping strategies of nurses at all levels of hospitals in Anhui Province during the COVID-19 outbreak, and to compare them with non-front-line prospective nurses (nursing college students).

## Methods

### Subjects

A questionnaire survey was conducted on online nurses and nursing college students in Anhui Province from February 1st to 9th, 2020, using a network questionnaire. A total of 850 questionnaires were sent, and 802 valid and complete questionnaires were recovered, with a recovery rate of 94.35%. Population composition: 202 males and 602 females; 298 participants in rural and 506 participants in urban; 374 nurses and 430 nursing college students; 377 participants from cities with severe epidemics (Hefei, Fuyang, Bengbu, the number of confirmed diagnoses is> 100), 170 participants from cities with moderate epidemic levels (Anqing, Wuhu, Lu’an, Suzhou, Ma’anshan, with confirmed diagnosis between 30-100), 257 participants from prefecture-level cities (Xuancheng, Huaibei, Huainan, Chizhou, Quzhou and Huangshan, the number of confirmed diagnoses <30).

### Research tools

#### Demographic information

It mainly includes the basic information of the participants, such as gender, age, identity, rural or urban, whether there is a confirmed or isolated person in the community or administrative village (in order to evaluate the spatial distance of the COVID-19 from the participants).

#### Emotional responses

Participants rated the extent that they experienced anxious, fear, sadness and anger in response to the outbreak of COVID-19 in An Hui on a 5-point scale, ranging from 1 (no such emotion) to 5 (the most intense feeling of the emotion). The order of presentation for these items was randomized.

#### Coping strategies

The tool for measuring the coping strategy during the outbreak of COVID-19 was revised based on the Brief COPE prepared by Carver (1997) [17,18]. Yeung and Fung (2013) used it to measure residents’ response strategies during the SARS outbreak which display a good reliability. The scale consists of problem-focused coping (active coping, planning, and use of instrumental support) and emotion-focused coping (use of emotional support, acceptance, positive reframing, religion, humor, substance use, self-distraction, self-blame, denial, behavior disengagement, and venting) consisting of 2 subscales with a total of 16 entries. We asked participants to report how often they used the strategy described in each project to respond to COVID-19, ranging from 1 (none) to 5 (always) on 5 levels. Higher scores indicated higher levels of coping. In this study, the Cronbach’s a coefficients of problem-focused and emotion-focused coping categories are 0.817 and 0.811 respectively.

### Statistical analyses

SPSS 21 software was used for data statistics. Independent sample t analysis, analysis of variance, correlation analysis and regression analysis were mainly used.

## Results

### General situations and difference of emotional responses and coping strategies

First, the independent sample t-test were conducted to examine participant identity, gender, urban-rural, and The severity of the urban covid-19 outbreak in each emotional responses, and the results are summarized in Table I. It was found that nurses’ anxiety (t_(799.33)_ = 3.05, *P* = 0.002), fear (t_(799.33)_ = 3.05, *P* = 0.002), sadness (t_(799.33)_ = 4.59, *P* = 0.000), and anger (t_(802)_ = 4.56, *P* = 0.002) was significantly higher than the emotional level of nursing college students. Women were significantly higher than men in terms of anxiety (t_(802)_ =-3.62, *P* = 0.000) and fear (t_(314.44)_ = 5.17, *P* = 0.000). The level of sadness (t_(584.85)_ =-3.85, *P* = 0.000) of participants from rural was significantly higher than that of participants from urban, while the anxiety of participants from urban (t_(679.52)_ = 2.55, *P* = 0.009) and anger (t_(802)_ = 3.04, *P* = 0.002) were significantly higher than participants in rural.

One-way ANOVAs were conducted to examine the severity of COVID-19 in the city in each emotion, and the results are summarized in Table I. In the result,no significant differences were found in anxiety 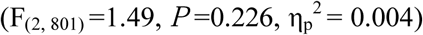, fear 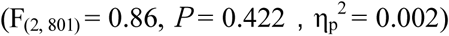, sadness 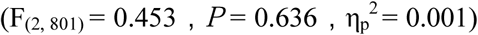, and anger 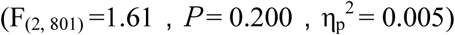.

One-way ANOVAs were conducted to examine the spatial distance of COVID-19 from the subject in each emotion, and the results are summarized in Table I. It was found that the main effects of fear 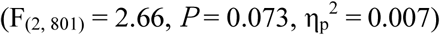 and sadness 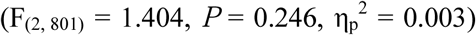 were not significant. A significant differences was found in anxiety 9.26, *P* = 0.000, 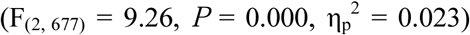, multiple comparisons found that anxiety in near space distance is significantly greater than far space distance (*P* = 0.000) and medium space distance (*P* = 0.073); the anxiety at medium space distance was significantly greater than that at far space distance (*P* = 0.008). The main effect of anger is aslo significant 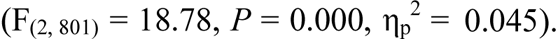 Multiple comparisons found that the anger emotion in near space distance is significantly greater than far space distance (*P* = 0.000) and medium space distance (*P*= 0.026), the anger in medium space distance is significantly greater than in far space distance (*P* = 0.000).

The independent sample t test found that nurses used problem-focused coping methods significantly higher than nursing college students (t_(802)_ = 4.99, *P* = 0.000). Women were significantly higher than men in problem-focused coping (t_(317)_ =-2.30, *P*= 0.022), and significantly lower than men in emotional-focused coping (t_(264.75)_ = 4.47, *P* = 0.000). There was no significant difference in the coping strategies between the participants from urban and rural.

One-way ANOVAs were conducted to examine the severity of COVID-19 in coping strategies, and the results are summarized in Table I. Three levels of severity did not differ in their use of problem-focused coping 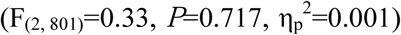 and emotion-focused coping 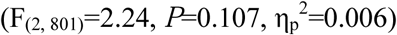toward COVID-19.

One-way ANOVAs were conducted to examine the spatial distance of COVID-19 from the subject in coping strategies, and the results are summarized in Table I. In the result, no significant was found in problem-focused coping 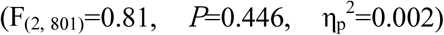 toward COVID-19. The main effect of emotion-focused coping was significant 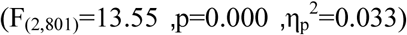, Multiple comparisons found that there was no significant difference in the emotion-focused coping between near space and far space distances (*P* = 0.715), and the emotion-focused coping at medium space distances were significantly higher than near space distances (*P* = 0.007) and far space distance (*P* = 0.000). That is to say, the level of emotion-focused coping at medium space distance is the highest, and the level of emotion-focused coping at far and near space distances is relatively lower.

### Correlation between emotional responses and coping strategies

After controlling for gender, spatial distance, identity, and urban-rural attributes, correlation analysis was performed on the emotional response and coping strategies. The results showed that there was a significant positive correlation between each four emotion (ps <0.001); a positive correlation was also found between problem-focused coping and emotion-focused coping (p <0.001). In addition to sadness, anxiety, fear, and anger were also positively correlated with Problem-focused coping and emotion-focused coping (ps <0.001).

### Regression analysis results of emotional responses and coping strategies

A stepwise multiple regression analysis was carried out to further reveal the correlations among emotion scores and coping strategies. In the regression analysis, emotion scores was used as the dependent variable, whereas two dimensions of coping strategies were used as independent variables (Table 3). Only problem-focused coping were included in the regression equation of anxiety, finding R^2^ = 0.035 and adjusted R^2^ = 0.034 (F_(1,802)_=29.17, p<0.000). This result demonstrates that the regression equation was significant, and problem-focused coping explain 3.4% of the variation. problem-focused coping and emotion-focused coping were included in the regression equation of fear, finding R^2^ = 0.063 and adjusted R^2^ = 0.060 (F_(2,801)_=26.79, p<0.000). This result demonstrates that the regression equation was significant, and two factor explain 6% of the variation. Only problem-focused coping were included in the regression equation of sadness, finding R^2^ = 0.01 and adjusted R^2^ = 0.009 (F_(1,802)_=8.25, p<0.004). This result demonstrates that the regression equation was significant, and problem-focused coping explain 0.9% of the variation. problem-focused coping and emotion-focused coping were included in the regression equation of anger, finding R^2^ = 0.029 and adjusted R^2^ = 0.026 (F_(2,801)_=11.77, p< 0.000). This result demonstrates that the regression equation was significant, and two factor explain 2.6% of the variation.

**Table 1:**
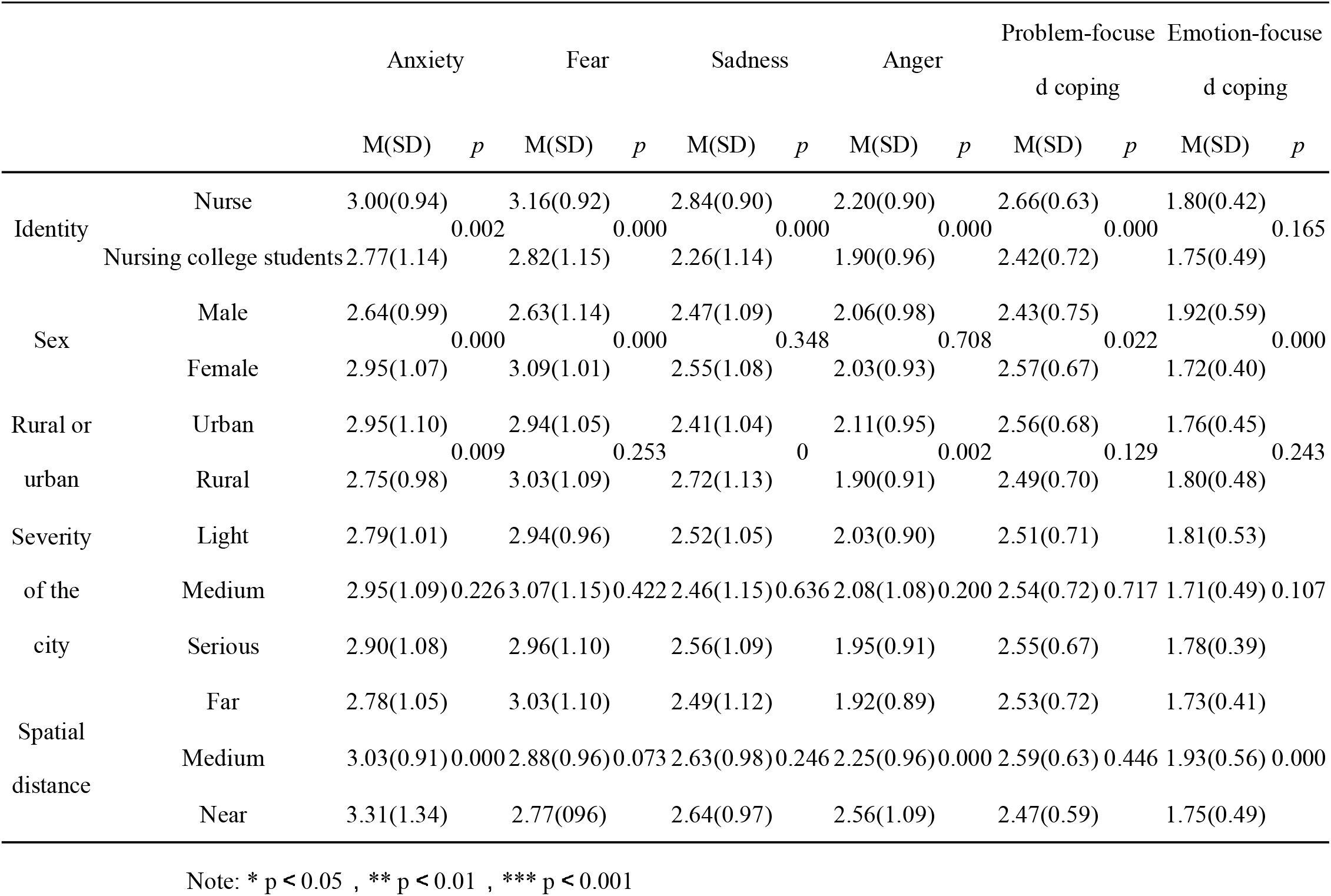
Descriptive statistics of emotion responses and coping strategies

|  |  | Anxiety |  | Fear |  | Sadness |  | Anger |  | Problem-focuse<br>d coping |  | Emotion-focuse<br>d coping |  |
| --- | --- | --- | --- | --- | --- | --- | --- | --- | --- | --- | --- | --- | --- |
|  |  | M(SD) | *p* | M(SD) | *p* | M(SD) | *p* | M(SD) | *p* | M(SD) | *p* | M(SD) | *p* |
| Identity | Nurse | 3.00(0.94) | 0.002 | 3.16(0.92) | 0.000 | 2.84(0.90) | 0.000 | 2.20(0.90) | 0.000 | 2.66(0.63) | 0.000 | 1.80(0.42) | 0.165 |
|  | Nursing college students | 2.77(1.14) |  | 2.82(1.15) |  | 2.26(1.14) |  | 1.90(0.96) |  | 2.42(0.72) |  | 1.75(0.49) |  |
| Sex | Male | 2.64(0.99) | 0.000 | 2.63(1.14) | 0.000 | 2.47(1.09) | 0.348 | 2.06(0.98) | 0.708 | 2.43(0.75) | 0.022 | 1.92(0.59) | 0.000 |
|  | Female | 2.95(1.07) |  | 3.09(1.01) |  | 2.55(1.08) |  | 2.03(0.93) |  | 2.57(0.67) |  | 1.72(0.40) |  |
| Rural or<br>urban | Urban | 2.95(1.10) | 0.009 | 2.94(1.05) | 0.253 | 2.41(1.04) | 0 | 2.11(0.95) | 0.002 | 2.56(0.68) | 0.129 | 1.76(0.45) | 0.243 |
|  | Rural | 2.75(0.98) |  | 3.03(1.09) |  | 2.72(1.13) |  | 1.90(0.91) |  | 2.49(0.70) |  | 1.80(0.48) |  |
| Severity<br>of the<br>city | Light | 2.79(1.01) |  | 2.94(0.96) |  | 2.52(1.05) |  | 2.03(0.90) |  | 2.51(0.71) |  | 1.81(0.53) |  |
|  | Medium | 2.95(1.09) | 0.226 | 3.07(1.15) | 0.422 | 2.46(1.15) | 0.636 | 2.08(1.08) | 0.200 | 2.54(0.72) | 0.717 | 1.71(0.49) | 0.107 |
|  | Serious | 2.90(1.08) |  | 2.96(1.10) |  | 2.56(1.09) |  | 1.95(0.91) |  | 2.55(0.67) |  | 1.78(0.39) |  |
| Spatial<br>distance | Far | 2.78(1.05) |  | 3.03(1.10) |  | 2.49(1.12) |  | 1.92(0.89) |  | 2.53(0.72) |  | 1.73(0.41) |  |
|  | Medium | 3.03(0.91) | 0.000 | 2.88(0.96) | 0.073 | 2.63(0.98) | 0.246 | 2.25(0.96) | 0.000 | 2.59(0.63) | 0.446 | 1.93(0.56) | 0.000 |
|  | Near | 3.31(1.34) |  | 2.77(0.96) |  | 2.64(0.97) |  | 2.56(1.09) |  | 2.47(0.59) |  | 1.75(0.49) |  |

Note: \* $p < 0.05$ , \*\* $p < 0.01$ , \*\*\* $p < 0.001$

**Table 2:**
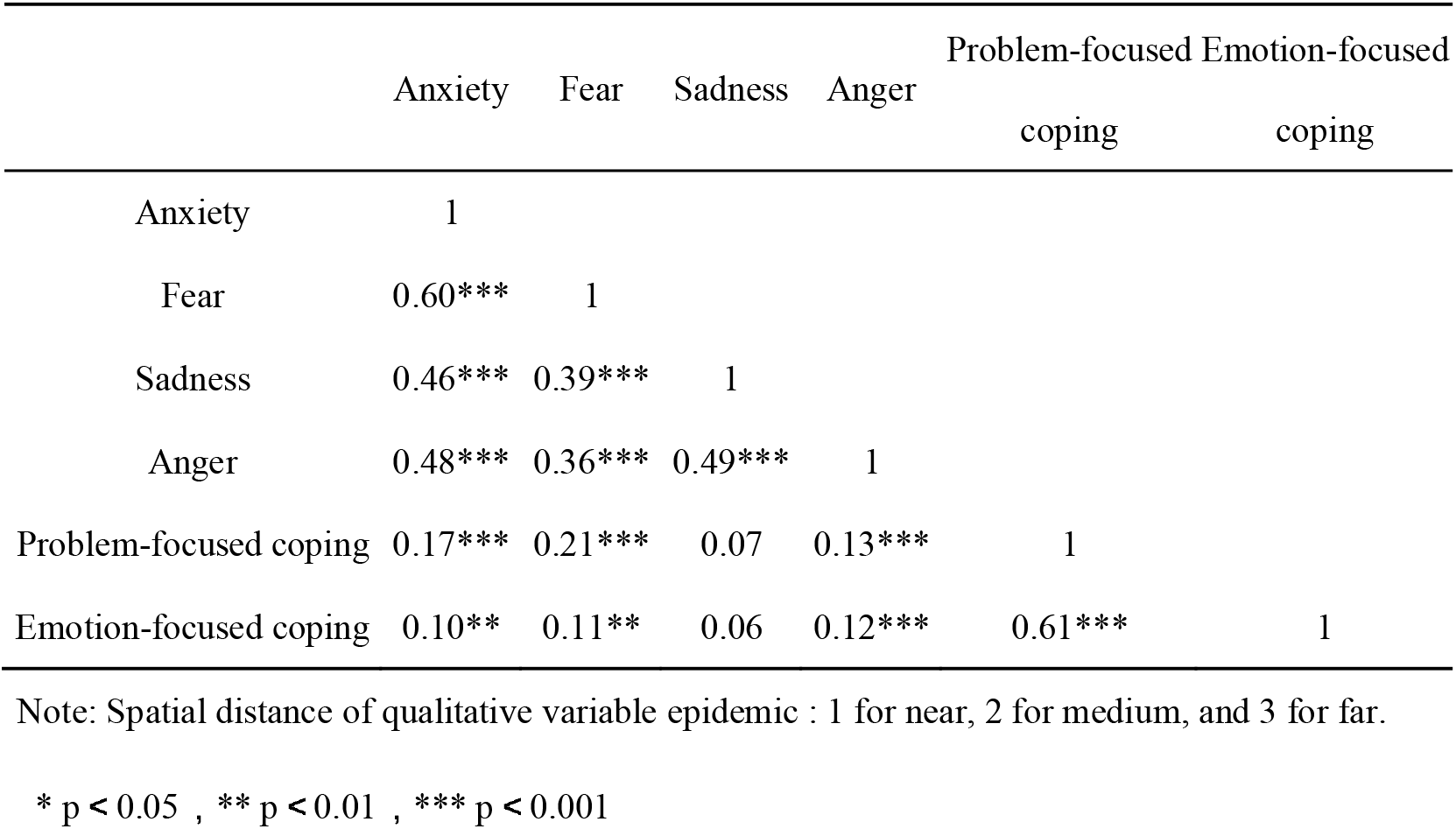
Correlations among emotion responses and coping strategies

**Table 3.**
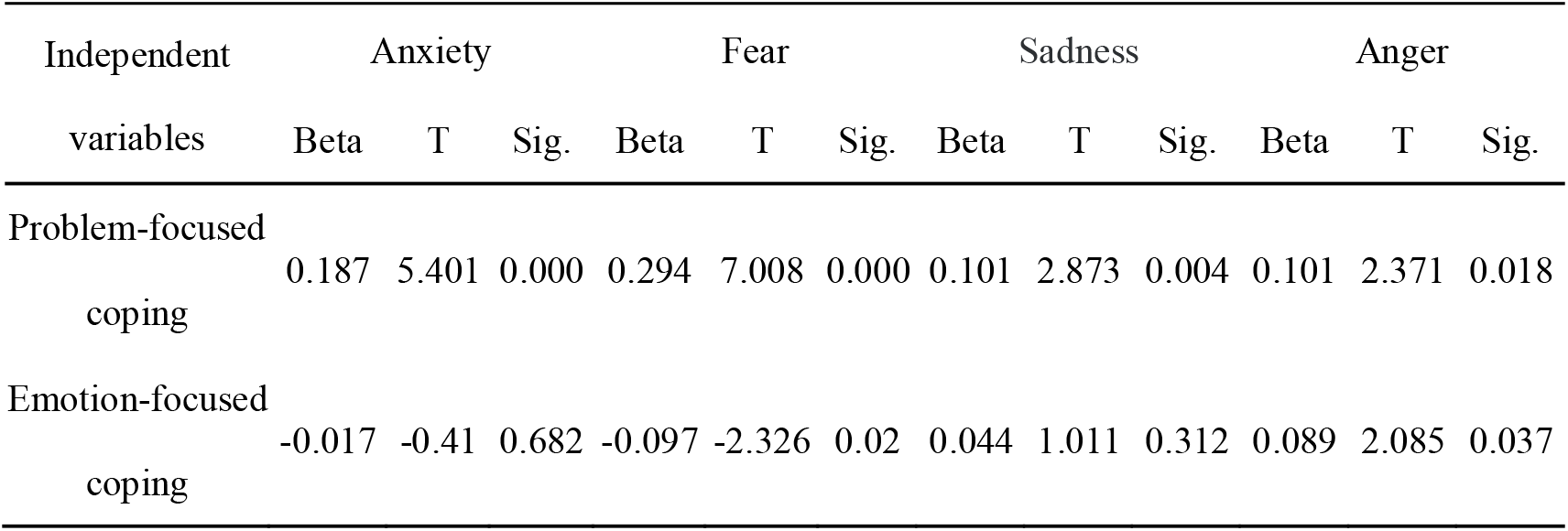
Regression analyses on emotion responses toward COVID-19 as a function of coping strategies.

A stepwise multiple regression analysis was carried out to further reveal the correlations among emotion scores and coping strategies. In the regression analysis, coping strategies was used as the dependent variable, whereas four dimensions of emotion scores were used as independent variables (Table 4). Only fear and anger were included in the regression equation of Problem-focused coping, finding R^2^ = 0.062 and adjusted R^2^ = 0.06 (F_(2,801)_ = 26.47, *P*<0.000). This result demonstrates that the regression equation was significant, and two factor explain 6% of the variation. Only anger were included in the regression equation of Emotion-focused coping, finding R^2^ = 0.023 and adjusted R^2^ = 0.022 (F_(1,802)_ = 19.11, *P*<0.000). This result demonstrates that the regression equation was significant, and the factor explain 2.2% of the variation.

**Table 4.**
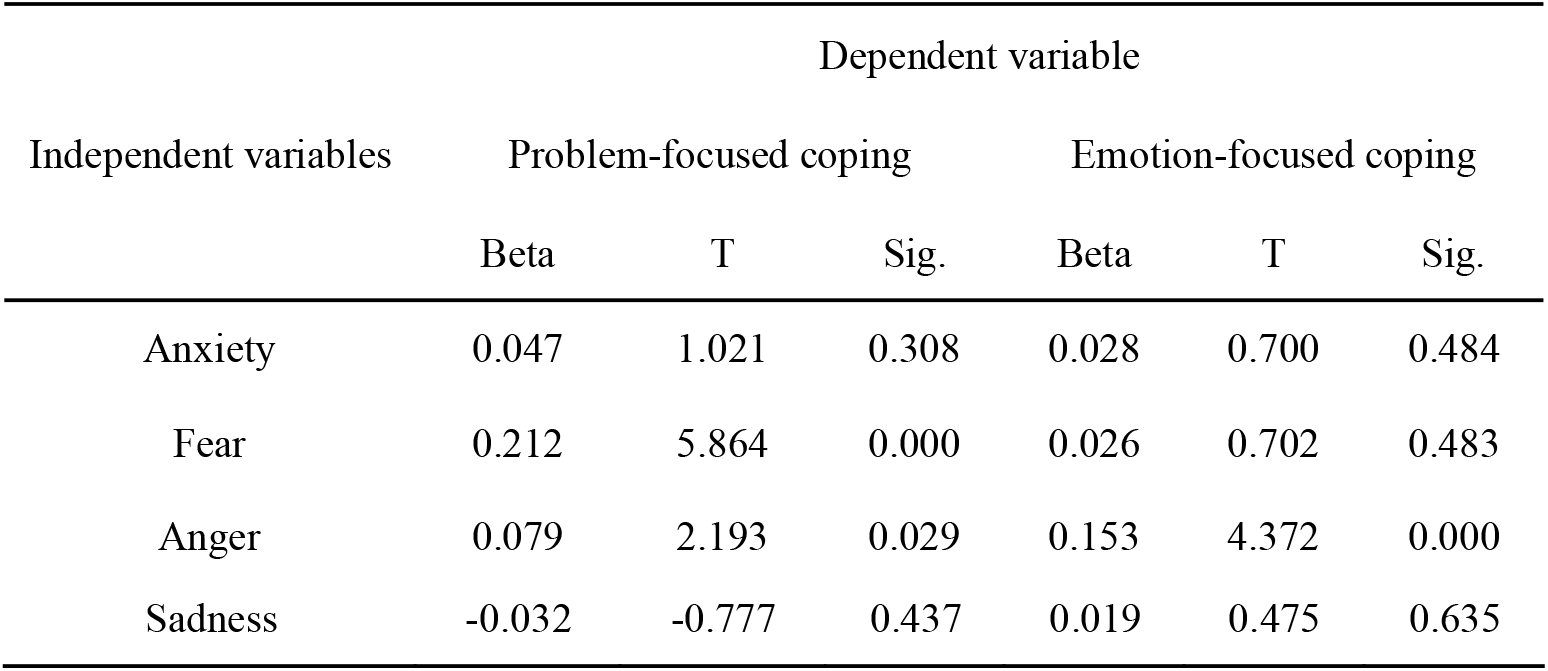
Regression analyses on coping strategies toward COVID-19 as a function of emotion responses.

## Discussions

COVID-19 is a stress source with great influence, both for individuals and social public groups. Different individuals may experience different levels of psychological crisis, especially those at the core of the incident. The study found that nurses at the heart of the event exhibit stronger anxiety, fear, sadness, and anger than nursing college students. Although, when encountering infectious disease incidents, nursing college students will also produce strong psychological stress and concerns about future careers, showing bad emotional and behavioral experiences of emotional excitement, doubt, and helplessness [19]. But nursing college students experience far less psychological stress than nurses. This has a lot to do with the working environment of nurses. Although nurses and nursing college students have similar professional knowledge, nurses, as one of the most vulnerable groups, are at the core of infection risk. They will be worried about being infected due to close contact with patients, unfamiliarity with special working environments and procedures, physical discomfort and inconvenience caused by special protection, Facing the suffering and death of critically ill patients, long-term separation from family members, and Risking their lives to live with patients every day, etc., which all cause a certain psychological response to medical staff. When he hear that his family is in trouble, he also feel sorry for being unable to do anything for his family. When seeing the patient is very distressed, and even if he has done his best to still not be able to save his life, he will also feel self-defeating psychologically, and think that he is not a good nurse, which resulting in strong self-blame and guilt. When a patient complains, he will feel aggrieved and not understood. Fighting against specific infectious diseases is a serious challenge for medical staff, especially for nurses, who are at risk of death at any time, which coupled with stressful work, sleep deprivation, low freedom, heavy responsibility, and high degree of cooperation. Due to the shortage of medical staff, nurses are faced with physical, mental and environmental stimuli, which leads to increased psychological load on nurses and more serious emotional problems [20]. This result is consistent with existing studies. This result is consistent with the results of a recent study by Xu and Zhang [21], who found that 85.37% (35 patients) of the first-line nurses fighting COVID-19 had adverse emotional reactions, including 2 nurses with depression, 16 nurses with anxiety, and 21 nurses with terror. During the SARS outbreak, many nurses showed conflicting roles as medical service providers and parents. On the one hand, they felt altruistic and professionally responsible. On the other hand, they were afraid and guilty that they might infect their families [22]. Nickell (2004)’s study found that about 20% of the population suffers from emotional depression during SARS, and the incidence of nurses is as high as 45% in Toronto [23].

Long-term research has generally found that women have significantly higher levels of depression, anxiety, and loneliness than men. This is considered to be related to gender traits. Women themselves attach more importance to their inner experiences and self-perceptions, their emotions are more fragile and sensitive, and they are more vulnerable to depression, anxiety and Loneliness [24]. Our study support this conclusion, which finds that women show more severe anxiety and fear than men. During the SARS outbreak, Gao et al. (2003) found that more women than men call for psychological counseling, and the content of the consultation is mainly emotional issues [24]. This indicates that there are gender differences in psychological characteristics in the face of public health emergencies. This study also found that participants from urban showed more anxiety and fear than participants from rural, but rural participants showed more sadness than urban participants. This may be that the city is densely populated and has a large flow of people, and the epidemic situation in urban is more serious. The COVID-19 is more closer to the participants urban than the rural participants. Urban participants are more concerned about whether they may be infected, so they are more anxious and fearful. On the contrary, rural participants pay more attention to the illness of others, and have sadness for patients. In addition, we found that the severity of the epidemic in the city has no difference in individual emotions, which may indicate that the individual is not very concerned about the severity of the epidemic in the city, or it may be caused by the small difference in the severity of the epidemic in the cities selected in our study. However, this study found that the more precise spatial distance between the epidemic and the participants significantly affected individual emotions. This study found that participants in the community or administrative village affected by COVID-19 (with diagnosed patients or isolators) had stronger anxiety and anger than those in the community or administrative village where were not affected by COVID-19. Participants in the community or administrative village with diagnosed patients reported significantly higher levels of anxiety and anger than participants in the community or administrative village with isolators. The presence of diagnosed patients in a community or administrative village means a higher probability of being infected. The geographical location of COVID-19 patients and the spatial distance of participants are a reflection of psychological distance. The closer the psychological distance is. The more people feel the danger, the more sexual and threatening, the more intense anxiety and anger. Our results and the research results during the outbreak of SARS support this conclusion, and this change is also in line with the psychological development law of people in response to stressful events [25].

College students often use immature or negative coping strategies instead of positive problem coping when faced with the pressure caused by public health emergencies [26]. As a medical worker with clinical work experience, the nurse has more knowledge and strategies to face similar pressures. This is consistent with our research, which found that nurses are more proactive in using problem-focused coping than nursing college students. This study also found that women are more likely to use problem-focused coping than men, and less likely to use emotional-focused coping. As mentioned earlier, women are more vulnerable and sensitive on emotional issues, so emotional-focused coping is rarely used when dealing with stress. In addition, this study also found that participants in the communities or administrative villages with isolators were more likely to using emotional-focused coping than those in the communities or administrative villages where was not affected or with diagnosed patients. This may be because when the communities or administrative villages is not affected by COVID-19, the participants do not pay enough attention to COVID-19 and do not cause a strong emotional response, so there is no need to use emotional-focused coping. When diagnosed patient (that is, the spatial distance is particularly close) appears in the community or administrative village, participants may not use sufficient emotional-focused coping due to the psychological typhoon eye effect [27].

In subsequent analysis, it was found that anxiety, fear, and anger were significantly positively related to problem-focused coping and emotion-focused coping. That is to say, there may be “the more coping the more panic” or “the more panic the more coping” or the “coping-panic cycle” The phenomenon. An analysis of the direction of the relationship between emotion and coping found that problem-focused coping has a significant predictive effect on anxiety and sadness. The explanatory power of anxiety is 3.4%, and the explanatory power of sadness is 0.9%, which indicating that the more problem-focused coping, the more anxious and sadness. Although both problem-focused coping and emotion-focused coping have significant predictive effects on fear, the total explanatory power is 6%, but the explanatory power of problem-focused coping is 5.6%, and the explanatory power of emotion-focused coping is only 0.4%. Therefore, the more problem-focused coping, the more fear. Problem-focused coping and emotion-focused coping both have a significant predictive effect on anger, with a total explanatory power of 2.6% and a problem-focused coping interpretation of 2.2%, but the explanatory power of emotion-focused coping is only 0.4%. This shows that problem-focused coping can predict emotional responses to a certain extent. This shows that there may be a phenomenon that “the more problem-focused coping, the more anxious, the more angry, the more sadness”.

Subsequently, our conducted a regression analysis on coping strategies, which found that fear and anger have a predictive effect on Problem-focused coping. The total explanatory power is 6%, the explanatory power of fear is 5.5%, and the explanatory power of anger is only 0.5%. Anger has a predictive effect on Emotion-focused coping, with an explanatory power of 2.2%. It can be seen that people may be the more fear, the more problem-focused coping, and the more angry, the more emotion-focused coping. Anxiety and sadness have no predictive effect on coping strategies. Combining the two parts of the regression analysis, it can be speculated that there may be a cycle of “the more fear—the more problem-focused coping” (forward interpretation power is 5.6%, reverse interpretation power is 5.5%), “the more angry, the more emotion-focused coping (2.2%) “,” The more problem-focused coping, the more anxious (3.4%), the more angry (2.2%), the more sadness (0.9%) “. During the outbreak of COVID-19, gender, urban/rural, and spatial distance will affect the anxiety, fear, sadness and anger and coping strategies of nurse and nursing college students. There is also a large emotional difference between nurses and nursing college students. Nurses have stronger emotional responses and are more willing to adopt Problem-focused coping. This study further reveals the direction of the relationship between emotional responses and coping strategies. In the outbreak of COVID-19, we suggest that hospitals should focus on providing psychological support to nurses, providing timely psychological assistance, training in coping strategies, providing adequate medical protective equipment, and taking a variety of interventions to block the spread of infectious diseases to form a medical environment where COVID-19 stops spreading in hospitals. It has created an optimistic environment and guarantees for personal safety for nurses, thereby enabling nurses to carry out ambulance work with high quality, and to provide a strong guarantee for resolutely winning the battle against epidemic prevention.

## Data Availability

The availability of all data referred to in the manuscript

## Ethical Approval

All procedures performed in studies involving human participants were in accordance with the ethical standards of the institutional and/or national research committee. The study was approved by the Academic Ethics Committee of Wannan medical college.

